# A Survey of Potential Bacterial Hazards in Salad Vegetables Sold in Lusaka District, Zambia, in 2018

**DOI:** 10.1101/2020.06.07.20124644

**Authors:** Chileshe Pule, Kunda Ndashe, Ruth Mfune, Abigail Chilumunda, Grace Mwanza, Bernadette Mumba, Lillian Mutesu Silavwe, Bernard Mudenda Hang’ombe

## Abstract

Salad vegetables are a source of fresh nutrition and can also serve a vehicle for foodborne pathogens. This was a cross sectional study conducted in February and March 2018 aimed at isolating some enteropathogens from some salad vegetables.

Ten samples of each salad vegetable (carrots, cabbage, cucumbers and tomatoes) were collected from two supermarkets and two open-markets. A total of 160 samples were collected and analysed microbiologically for presence of *Shigella* spp, *Vibrio* spp and *Salmonella* spp. The isolated bacteria were identified by conventional biochemical tests.

*Vibrio* spp were isolated from all the vegetables that were collected from both the two open markets (62.5% and 72.5%) and two supermarkets (67.5% and 77.5%), with the highest isolation rate recorded in cabbages. The frequency of occurrence of *Shigella* spp and *Salmonella* spp was higher in supermarkets (50% and 65%; 50% and 15%) than in the open markets (10% and 7.5%; 10% and 35%), respectively.

The present study revealed the presence of potential bacterial hazards in salad vegetables sold in both open markets and supermarkets in Lusaka district. The isolated bacterial pathogens are aetiological agents of diarrheal disease that even at low dose could be infectious for sensitive populations.

## 1. Introduction

In the quest for healthy life styles there has been an increase in the consumption of vegetable salads which are normally eaten without any heat treatment and sometimes with minimal cleaning with water. Salad vegetables include carrots (*Daucus carota*), cabbage (*Brassica oleracea*), cucumbers (*Cucumis sativus*) and tomatoes (*Solanum lycopers*). Researchers have demonstrated that vegetables may serve as vehicles for transmission of enteric bacteria that are responsible for foodborne illnesses [1-3]. Developed countries such as the United States and European Union reported a total of 377 and 198 fresh produce-associated outbreaks, respectively, between 2004 and 2012 [4]. Foodborne illness is a major public health concern worldwide in terms of numbers of persons affected and economic cost. Approximately one in 10 people in the world fall ill food and 420 000 die every year after eating contaminated food [5].

Zambia continues to face immense challenges with foodborne and waterborne illness. In 2017, the country experienced an outbreak of cholera in Lusaka with 3938 cases and 82 deaths (case fatality rate, 2.1%) [6]. The main source of transmission during the outbreak was contaminated water. Furthermore, several studies conducted in Lusaka have reported that many water sources such as municipal and ground water are contaminated with faecal coliforms both in urban and peri-urban townships [7-9]. Vegetables easily become contaminated with pathogenic microorganisms from farm to fork and water used for irrigation and processing post-harvest contributes to the bacterial quality of vegetables [10]. Unsafe water used for rinsing the salad vegetables before eating and for sprinkling at retail points to keep them fresh is also a source of contamination [11]. Microorganisms like *Escherichia coli, Salmonella* spp, *Klebsiella* spp, *Shigella* spp and other coliforms may pose serious health threats to consumers [12].

In Lusaka, local farmers use municipal and groundwater for irrigation and washing of salad vegetables post harvest. The bacteriological quality of the water is reported to be poor and therefore can serve a as source of contamination. To the best of our knowledge there exists no publication reporting the bacteriological quality of salad vegetables in Lusaka. Therefore, we conducted this study to investigate the presence of faecal bacteria (*Salmonella* spp, *Shigella* spp and *Vibrio* spp) on salad vegetables in retail sale points such as supermarkets and open markets of Lusaka district.

## 2. Material and Methods

### Sampling and sample preparation

The present cross-sectional study was carried out during a 2-month period from February through March 2018. Salad vegetables were collected from two supermarkets and two open markets and were examined at the University of Zambia, School of Veterinary Medicine Bacteriology laboratory. The two supermarkets (A and B) and the open markets (Soweto and Mtendere) were included in the study because they are the main sources of fresh produce in Lusaka. The vegetables that were collected included carrots (*Daucus carota*), cabbage (*Brassica oleracea*), cucumbers (*Cucumis sativus*) and tomatoes (*Solanum lycopers*).

Convenience samples were taken and the sample size was limited by the cost of the laboratory testing, a total of 160 samples were collected for the study. Forty salad vegetables, 10 of each type (carrots, cabbage, cucumbers and tomatoes) were randomly collected from each participating supermarkets or open markets. From the two open markets, 8 vegetable traders (4 from each) were randomly selected from whom one type of salad vegetable was sampled. Each sample was secured in a sterile plastic bag and then transported in cooler boxes surrounded by ice packs to the laboratory, where they were analysed for potential bacterial hazards with 2 hours of collection.

The study was conducted after ethical approval from ERES CONVERGE IRB, and consent was sought from the management of the participating supermarkets and vegetable traders in the open markets.

### Microbiological analysis

An estimated 25 grams of each sample were weighed aseptically in a sterile test tube and were vortexed with 225ml of buffered peptone water (BPW) for 2 minutes. The mixture was kept for 1 hour at room temperature before analysis. Salmonella spp. were detected in four successive steps. Pre-enrichment in BPW at 37 ° C for 20 hours, followed by enrichment in selenite broth incubated at 37 ° C for 24 hours. The isolation was done on selective media xylose lysine deoxycholate (XLD) agar at 37 ° C for 24 hours. Suspect *Salmonella* colonies (red colonies with black centres) were confirmed with conventional biochemical tests. Typical *Salmonella* isolates were further serotyped by using using *Salmonella* O and H agglutination antisera following the Kauffman-White serotyping scheme [14].

For detecting the presence of *Shigella* spp, 25 g portions of each item were enriched in a 225 ml *Shigella* broth (Difco, Detroit, MI, US) containing novobiocin, for 22 hours at 42°C. Enriched cultures were streaked onto MacConkey agar (Himedia, India) and incubated at 35°C for 20 hours. Presumptive *Shigella* isolates (slightly pink and translucent) were identified by conventional biochemical methods.

For the detection of *Vibrio* spp, 25g of each sample was enriched in 225ml of alkaline peptone water (Himedia, India) and incubated at 37 °C for 6 hours. The enriched cultures were streaked on thiosulfate citrate bile sucrose (TCBS) agar (Himedia, India) followed by incubation al 37 °C for 24 hours. Suspected vibrio colonies (yellow and green) were further identified by conventional biochemical methods.

## 3. Results

The presence of *Vibrio* spp, *Shigella* spp and *Salmonella* spp in the samples was identified. The Vibrios were isolated from all the salad vegetables that were collected from the open markets (62.5% and 72.5%) and supermarkets (67.5% and 77.5%) as shown in the table. The rate of isolation of *Vibrio* spp in each type of vegetables varied, lowest and highest isolation rate of the Vibrios was recorded in cucumber and cabbages, respectively.

*Shigella* spp were not isolated from tomatoes and cucumbers from open markets and the highest isolation rate, 70% and 80%, of the bacterium was recorded in tomatoes collected from supermarkets 1 and 2, respectively. *Shigella* spp were isolated from all the vegetables in supermarket 1 and 2 (50% and 65%) while the bacterium was isolated from cabbages and carrots from open market 1 and 2 (10% and 7.5%) as shown in the table.

*Salmonella* spp isolation rate was highest in vegetables in supermarket 1 and open market 2 (50% and 35%). Cabbages and carrots from Supermarket 1 and tomatoes from open market 1 recorded the highest isolation at 70%. *Salmonella* spp were not isolated from carrots in open market 2 and supermarket 2, cucumbers and cabbages from open market 1.

## 4. Discussion

The safety of salad vegetables is based on the presence and quantities of pathogenic microorganisms they contain. This study aimed at isolating some diarrheagenic bacterial pathogens in readily available salad vegetables. The study reported the presence of *Vibrio* spp, *Shigella* spp and *Salmonella* spp contamination of the commonly consumed salad vegetables in Lusaka district. Studies from Togo, Iran, Hong Kong, Saudi Arabia and Nigeria demonstrated the presence of coliforms and other diarrheagenic bacteria in fresh cut and salad vegetables [15-18].

From the present study, *Vibrio* spp was isolated from all the salad vegetables except carrots collected from the open markets. The frequency of occurrence of the bacterium in vegetables from open markets 1 and 2 (72.5% and 62.5%) was similar to those recorded from supermarket 1 and 2 (67.5% and 77.5%). This result differs markedly from those reported in Malaysia where vegetables from open market had a higher frequency of occurrence of *Vibrio* spp than those from supermarkets [19]. In a study by Tunung and colleagues (2010), *V. parahaemolyticus* was isolated from 20.65% of salad vegetables collected from all retail outlets in Selangor, Malaysia [19]. In Bangladeshi, Rahman and Noor (2012) reported the presence of *Vibrio spp* in cucumbers, carrots, lettuce and tomatoes, which were only isolated after enrichment [20]. *Vibrio* spp are waterborne and therefore irrigation of vegetables with waste water or organic manure may serve as a source of contamination. In a study by Okafo and colleagues (2003) in Kowa and Sabon Garri, Nigeria, they reported high coliform counts in both irrigation water and vegetables and they further recorded higher frequency of isolation of *Vibrio* spp in the dry season [21]. The present study coincided with the 2017/2018 cholera outbreak in Lusaka District but it cannot be concluded that the *Vibrio* spp isolated from the vegetables contributed to the transmission of the disease as it was not identified as *V. cholerae* O1/O139.

The frequency of occurrence of *Shigella* spp and *Salmonella* spp from open market 1 and 2 (10% and 7.5%; 25% and 35%) was lower than supermarket 1 and 2 (50% and 65%; 50% and 15%), respectively. This result does not conform with common trend in other studies where open markets report more bacterial contamination of vegetables when compared to supermarkets. From the open markets *Shigella* spp were only isolated from cabbages and carrots, while the bacterium was isolated from all the vegetables sampled in the supermarkets. The higher rate of isolation of *Shigella* spp and *Salmonella* spp from the supermarkets could be attributed to storage duration of the vegetables and possibilities of cross contamination between the products as well. The environmental temperatures in the supermarkets are not low enough to prevent the proliferation of mesophiles. Furthermore, the generation time of *Salmonella* spp and *Shigella* spp is between 34 to 40 minutes and therefore storage of vegetables for more than 48hrs at environmental temperature (24 to 29 °C) will result in multiplication of the bacteria to infectious doses. In a study by Silva and colleagues (2009), *S. enterica subsp. enterica sorovar Typhimurium* stored at 25 °C for 24hrs increased to population of 5 to 6 log10 at generation time of 34.8 minutes [22]. The study highlighted that at environmental temperature (24 to 29 °C) *Salmonella* thrives well and therefore explaining the high isolation rates in supermarkets were the vegetables are stored for more than two days. At open markets, the traders normally buy the vegetables daily and sell them off during one work shift, and therefore they are not stored overnight.

The presence of diarrheagenic bacterial pathogens in vegetables from all retail points in the study suggests that contamination could have occurred prior to and post harvesting and during storage in the sales outlets. Other researchers have highlighted that the main sources of contamination of vegetables with enterobacteriaceae include irrigation with waste water, application of contaminated organic fertilizers, washing with contaminated water, and handling by infected food handlers and vendors [23-27]. For pathogens with no animal reservoir, such as *Shigella* spp., human faeces provide the primary source of contamination, and infected food handlers have frequently been implicated as the cause of outbreaks [28]. The high isolation rate of the enteropathogens in the study suggest high bacterial contamination of the salad vegetables which is a reflection of storage condition and how long these vegetables were kept before they were obtained for sampling. Although the quantity of the enteropathogens in the vegetables, colony forming unit per gram (cfu/g), was not determined, the presence of these enterobacteriaceae is of public health concern.

## 5. Conclusion

The present study revealed the presence of a potential bacterial hazard on salad vegetables sold in Lusaka district. The isolated bacteria pose a public health risk considering that the salad vegetables are consumed without heat treatment and with minimal washing. Vegetable salads are a significant part of urban food supplies and therefore adequate measures ought to be taken to reduce the microbial load on the vegetables. Intervention by the local authority (Lusaka City Council) and the Ministry of Agriculture is required by engaging farmers and salad vegetable traders in hygiene practices and responsible farming of vegetables to prevent pre and post harvest contamination. Furthermore, the general public should thoroughly clean salad vegetables with clean water before consumption.

## Data Availability

The authors confirm that the data supporting the findings of this study are available within the article

## Acknowledgement

We would like to recognize that this was a study undertaken by Ms. Chileshe Pule as partial fulfilment for Bachelor of Science Environmental Health of Lusaka Apex Medical University (LAMU). We wish to express our gratitude to the staff at the University of Zambia, School of Veterinary Medicine, Bacteriology Laboratory for technical help rendered during bench work sessions of the research. We would like to thank the academic staff of LAMU, Faculty of Health Sciences for the support and contribution towards the research. Finally, we are indebted to the market traders of Soweto and Mtendere markets that allowed us to conduct the research in their vegetable to them we say thank you.

